# A framework for detecting causal effects of risk factors at an individual level based on principles of Mendelian randomization: Applications to modelling individualized effects of lipids on coronary artery disease

**DOI:** 10.1101/2024.01.18.24301507

**Authors:** Yujia Shi, Yong Xiang, Yuxin Ye, Tingwei He, Pak-Chung Sham, Hon-Cheong So

**Author notes:** Corresponding author: Hon-Cheong SO.

## Abstract

Mendelian Randomization (MR), a method that employs genetic variants as instruments for causal inference, has gained popularity in assessing the causal effects of risk factors. However, almost all MR studies primarily concentrate on the population’s *average* causal effects. With the advent of precision medicine, the *individualized* treatment effect (ITE) is often of greater interest. For instance, certain risk factors may pose a higher risk to some individuals compared to others, and the benefits of a treatment may vary among individuals. This highlights the importance of considering individual differences in risk and treatment response.

We propose a new framework that expands the concept of MR to investigate *individualized* causal effects. We presented several approaches for estimating Individualized Treatment Effects (ITEs) within this MR framework, primarily grounded on the principles of the”R-learner”. To evaluate the existence of causal effect heterogeneity, we proposed two permutation testing methods. We employed Polygenic Risk Scores (PRS) as the instrument and demonstrated that the removal of potentially pleiotropic SNPs could enhance the accuracy of ITE estimates. The validity of our approach was substantiated through comprehensive simulations.

We applied our framework to study the individualized causal effect of various lipid traits, including Low-Density Lipoprotein Cholesterol (LDL-C), High-Density Lipoprotein Cholesterol (HDL-C), Triglycerides (TG), and Total Cholesterol (TC), on the risk of Coronary Artery Disease (CAD) using data from the UK Biobank. Our findings indicate that an elevated level of LDL-C is causally linked to increased CAD risks, with the effect demonstrating significant heterogeneity. Similar results were observed for TC. We also revealed clinical factors contributing to the heterogeneity of ITE based on Shapley value analysis. Furthermore, we identified clinical factors contributing to the heterogeneity of ITEs through Shapley value analysis. This underscores the importance of individualized treatment plans in managing CAD risks.

## 1 Introduction

The rising incidence and mortality rates of chronic diseases have imposed a significant burden on numerous countries over the past decades^1^. Consequently, identifying potential causal risk factors and designing appropriate interventions have emerged as top priorities. In the past, epidemiologists focused primarily on studying the average causal effect of prospective interventions, thereby overlooking the importance of population heterogeneity. The presence of heterogeneity suggests that individuals may derive varying benefits from the same intervention. For instance, a randomized controlled trial (RCT) demonstrated that metformin could have a heterogeneous impact on diabetes mellitus (DM) prevention among patients with impaired glucose metabolism^2,3^. Specifically, patients at a higher risk of diabetes might experience a more substantial absolute risk reduction than those at lower risk. This study underscores the importance of estimating individualized treatment effects (ITEs). To optimize intervention efficiency across the population and minimize costs, it is important to estimate the potential benefit a specific patient may gain from an intervention (or risk factor prevention). In this study, we aimed to estimate the individualized causal treatment effect of a given intervention to individual patients, leveraging the principles of Mendelian randomization (MR).

It is widely accepted that the most accurate approach to estimate the causal effect is via a randomized controlled trial (RCT), in which both known and unknown confounding factors can be controlled for by treatment randomization^4^. However, RCTs are often prohibitively expensive, limited by ethical considerations or logistical constraints, and may lack generalizability due to strict inclusion/exclusion criteria^5,6^. Consequently, researchers frequently resort to observational studies to estimate causal effects. Unlike RCTs, a major concern with observational studies is that unmeasured confounding may influence causal inference. Mendelian Randomization (MR) serves as a valuable approach to mitigate the risk of unmeasured confounding and is largely immune to reverse causality. In MR, genetic variants are utilized as instruments to represent the exposure^7^.

Following years of development and innovation, a variety of statistical methods have been established for Mendelian Randomization (MR) analyses, including the Wald ratio method, two-stage least squares, MR-Egger, weighted median, and semi-parametric methods^8^. Although these methods are robust and flexible, they still have limitations. An important one is that they can only estimate an *average* causal effect without considering the heterogeneity of the population, and there is a lack of innovations regarding the estimation of *individualized* treatment (or risk factor) effects.

Our main contribution is the introduction of a novel framework, MR-ITE, capable of inferring individualized causal effects in observational studies based on the MR approach, utilizing the polygenic risk score (PRS) as an instrument. We have proposed several Individualized Treatment Effect (ITE) estimation methodologies within the MR framework, grounded on the principles of “R-learners”^9^. These methods offer high flexibility as they leverage supervised machine learning (ML) approaches for modelling, imposing virtually no restrictions on the type of ML models employed. Our other contributions to the MR-ITE framework include: (1) To mitigate the risks of bias from invalid instruments, we suggest the use of the contamination mixture approach to eliminate potential pleiotropic Single Nucleotide Polymorphisms (SNPs) prior to calculating the PRS^10^; (2) We also presented permutation-based approaches to test for the presence of heterogeneity under MR-ITE; (3) We proposed methods to evaluate the most important effect modifiers in MR-ITE analyses, for instance, by employing the Shapley value; (4) We applied our proposed framework to study the individualized effects of lipids on risks of coronary artery disease (CAD); Our findings indicate that LDL-C and TC may exert heterogeneous causal effects on CAD risks, and we have identified major effect modifiers. In conclusion, this work underscores the importance of individualized causal effect estimation in observational studies and presents innovative methodologies to achieve this goal.

## 2 Methods

### 2.1 Set-up and notation

#### 2.1.1 Rubin’s causal model

A causal model needs to be formalized first. A well-established and popular choice is the Neyman-Rubin causal model, also called the potential outcome (counterfactual) framework^11^. We consider a dataset with *N* units, indexed by *i* = 1, …, *N*. Following the potential outcome framework, we define the potential outcome for unit *i* in treatment and control status as *Y*_*i*1_ and *Y*_*i*0_, respectively. For each unit, we let *X*_*i*_ be a vector consisting of *M* covariates and *Z*_*i*_ be a continuous instrument variable. We further define *T*_*i*_ ∈ {0,1} as a binary indicator for the treatment, where *T*_*i*_ = 0 means that the unit *i* does not receive any treatment and *T*_*i*_ = 1 means that the unit *i* is receiving the treatment. Given the formalization above, our data can be regarded as a set of quadruple data point 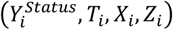 units, indexed from 1 to *N*. In this case, we further define the unit-level causal effect as the difference between two potential outcomes *Y*_*i*1_ and *Y*_*i*0_, τ_*i*_ = *Y*_*i*1_ −*Y*_*i*0_.

The framework discussed above is under a binary treatment setting. However, in many epidemiology studies, the risk factors are continuous variables, and it may be difficult to define an arbitrary cutoff to partition the population into treatment and control groups. The main difference between binary and continuous settings is that we no longer define a binary indicator for the treatment. In this case, the unit-level causal effect is defined as the effect of unit increment of treatment on the outcome, 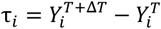

#### 2.1.2 Assumptions of Instrumental Variables

As we regard our approach as an extension of the Mendelian Randomization framework, we also require similar assumptions that the MR framework needs to achieve a consistent estimation of the causal effect. A natural way to extend the core assumptions for MR to individualized MR applying PRS as the instrument as follows:

1. The polygenic risk score is associated with the exposure.
2. The polygenic risk score is independent of the outcome given the exposure and possible confounders (measured and unmeasured) of the exposure-outcome association.
3. The polygenic risk score is independent of factors that confound the exposure-outcome relationship.

The first assumption is a key assumption of instrument analysis that we require the instrument to have a non-zero effect on the treatment, at least for some covariates, referring to the instrumentation assumption or relevance assumption. The second assumption is the exclusion restriction assumption, which requires the instrument only to affect the outcome through its effect on the exposure or treatment. The last assumption is similar to the second one, where the instrument should not have effect on the outcome through the confounders^12,13^.

### 2.2 Generalized Random Forest (GRF), including GRF using instrumental variables (IV-GRF)

The generalized random forest (GRF) is a forest-based estimator introduced by Athey et al^14^. It preserves the main components of the classic random forest, including recursive partitioning and feature subsampling^14^. For details of the methodology and notations, please refer to^14^. Briefly, GRF can be considered as an adaptive nearest neighbor with weights *α*_*i*_(*x*) that can be used in local estimating equations 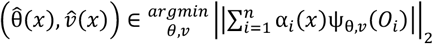 where ψ(⋅) is some scoring function and *v*(*x*) is the optional nuisance parameter.

We can think of the weight *α*_*i*_(*x*) as a measurement of the relevance between the *i*th training sample and a given sample *x* in fitting *θ*(⋅), the function returning the individualized treatment effects (ITE). GRF uses the frequency that the *i*th training sample and the given sample *x* are located in the same leaf to measure such relevance. It can be defined as 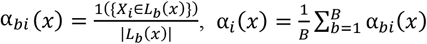 where *B* denotes the number of trees in the forest and *b* denotes the *b*^th^ tree. *L*_*b*_(*x*) is the set of training examples falling in the same “leaf” as *x*.

The instrumental causal forest (IV-GRF) is an important application of GRF, in which the gradient-based labelling ρ_*i*_ is modified to fit the instrumental variable case. In the regular GRF causal forest, the gradient-based labeling is defined as, 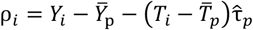 while in the iinstrumental causal forest, the gradient-based labeling is defined as, 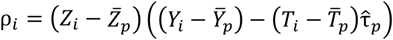 where the 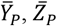 and 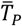 stand for the average of *Y, Z* and *T* over the parent node *P*. The algorithm will further run a standard CART regression split by splitting the parent node into two child nodes *C*_1_ and *C*_2_ with maximizing the criterion:

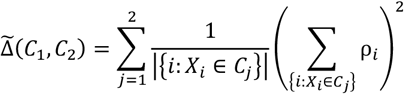

In addition to the difference in making splits, the instrumental GRF modifies the prediction step by introducing the forest-weighted two-stage least squares approach, allowing a more accurate causal effect estimation in the instrumental case.

### 2.4 Double robustness instrumental variable estimator (DRIV)

Unlike instrumental causal forest, DRIV mainly focuses on designing a novel loss function based on machine learning models^16^. The method starts by a ‘preliminary’ estimate of the individualized treatment effect (ITE), which can be computed by minimizing the following loss function:

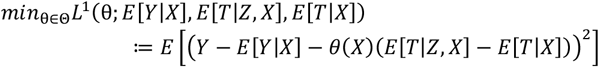

where *Z* is the instrumental variable and *θ* represents estimate of the ITE. The above estimate is referred to as the double machine learning IV (DMLIV) estimate as described in ref ^16^.

The nuisance terms *E*[*Y*|*X*],*E*[*T*|*Z,X*] and *E*[*T*|*X*] can be estimated from an independent training set using any supervised learning models. In our application, we applied the DRIV algorithm implemented by EconML package to estimate the ITEs^17^. Since finding a separate dataset is usually difficult, the above terms are estimated based on cross-validation (CV) to avoid overfitting, following EconML’s implementation. Also following the default settings by EconML, we used WeightedLassoCV to model the continuous dependent variables and used the LogisticRegressionCV to model the binary ones. Both WeightedLassoCV and LogisticRegressionCV are originally implemented in scikit-learn package^18^. The LogisticRegressionCV class implements a logistic regression with L2 penalty, with hyperparameters determined by cross validation. WeightedLassoCV is a LASSO linear model with hyper-parameters determined by CV. However, we emphasize that *any* supervised ML models can be used to estimate the above terms.

However, the 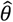 estimated from the first step was shown to be not robust enough^16^. An improved estimator that is doubly robust can be applied instead^16^,

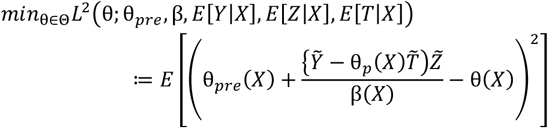

where β(X) =*E*[(*T* − *E*[*T*|*X*])(*Z* − *E*[*Z*|*X*])|*X*]. This term is also estimated by CV similar to above.

Note that we can further apply SHAP analysis directly to assess the effect modifier importance since the DRIV allows a flexible specification of machine learning models in the training step, which is not feasible in a causal forest model. SHAP can only provide an approximation of variable importance of causal forest model, as causal forest does not make the predictions based on a simple averaging of predictions of individual trees^14,19^.

### 2.5 Polygenic Risk Score

A single trait-associated genetic variant can usually only explain a small proportion of variance of the phenotype. Evidence suggests that many common disease/phenotypes are mediated by multiple genetic variants simultaneously, where each individual genetic variant only contributes a small effect^20,21^. Polygenic risk score can be used to summarize the estimated effect of multiple trait-associated genetic variants on an individual’s phenotype, which is usually defined as a weighted sum of trait-associated risk alleles across multiple genetic loci^22^.

In our study, we apply PRSice-2 to calculate the individualized PRS as an instrument for further analysis. PRSice-2 is an efficient PRS calculating software that can automate PRS analyses, which can identify the precise P-value threshold by calculating the PRS at multiple threshold^23^. In our analysis, we set the P-value threshold to 5e-8.

### 2.6 Contamination Mixture Model

A major concern of applying an allelic score as an instrument in MR is that it may violate the assumption of an instrumental variable. Including pleiotropic SNPs in calculating the allelic score may significantly affect the overall performance of the results^24^. Therefore, it is necessary to remove the pleiotropic SNPs to ensure the calculated PRS is a valid instrument. Here we assume that same SNPs show imbalanced horizontal pleiotropy both at the population and individual level. In this study, we propose the application of the contamination mixture (ConMix) approach to identify invalid SNPs in our framework^10^. This approach has been shown to be robust to the presence of pleiotropic effects^25^. The contamination mixture approach assumes that the valid genetic variants follow a normal distribution centered at the true causal parameter θ with standard deviation equal to the ratio estimate’s standard error 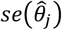 while the invalid genetic variant is normally distributed around 0, with variance comprising the uncertainty in the ratio estimates (which is 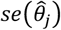) and the proposed variability in the invalid estimand (denoted as ψ^2^). The ConMix approach first specify a likelihood function *L*(*θ*, ξ) integrating information from all genetic variants, defined as,

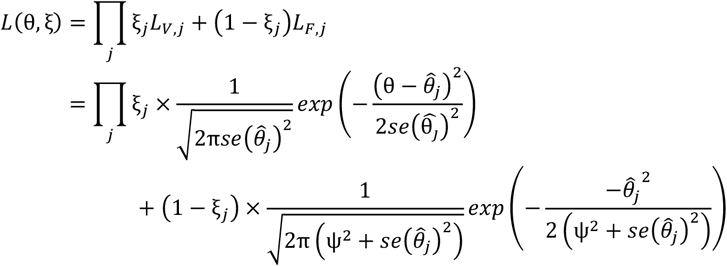

where ξ is a vector denoting the configuration of the valid and invalid instruments, *L*_{*V,j*}_ is the likelihood contribution when genetic variant *j* is valid and *L*_{*F,j*}_ is the likelihood contribution when genetic variant *j* is invalid. In the likelihood, ξ_*j*_ = 1 if genetic variant *j* is a valid instrument, while ξ_*j*_ = 0 if genetic variant *j* is invalid. Instead of making inferences with this likelihood, the ConMix method applies a profile likelihood approach, in which the causal estimate *θ* is assumed fixed. When *θ* is fixed, the optimal *θ* that maximizes the aforementioned likelihood function can be easily inferred, that is, ξ_*j*_ equals to 1 when *L*_*V,j*_ is greater than *L*_*F,j*_ for genetic variant *j*. The ConMix approach iterates through a range of values of *θ* and picks the one maximizing the profile likelihood. In summary, the ConMix approach tries to identify the invalid SNPs based on the distance between variant-specific estimate 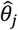 and the proposed causal parameter *θ*, in other words, if the 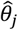 is closer to the *θ*, the SNP *j* is more likely to be a valid SNP. Here we removed the likely pleiotropic SNPs based on the ConMix approach before construction of the PRS as the instrument.

### 2.7 Assessing the Presence of Treatment Effect Heterogeneity

In addition to estimating the individual treatment effect, we also presented two permutation-based methods to assess the presence of heterogeneity among the estimated treatment effect. Typically, the heterogeneity of treatment effect can be defined as a non-random, explainable variability in the direction and magnitude of individualized treatment effect obtained from a population^26^. Another way to think of heterogeneity of treatment effects is that whether the predicted treatment effects are different from the average effect by chance^27^. These definitions offer valuable insight in developing methods for us to estimate ITE and assess heterogeneity. For example, in each split, the causal tree will try to maximize the variance of the estimated treatment effect across leaves as well as penalize the uncertainty of estimated treatment effects^28^. If covariates cannot contribute to the heterogeneity, the variance of the predicted ITE across leaves should be smaller than splitting on covariates that contribute to the heterogeneity, inspiring us to develop the heterogeneity testing methods based on covariate permutation.

Here we present two permutation-based methods, including the permutation-variance and permutation-τ-risk test, to evaluate whether there is significant heterogeneity. While the same principles can also be applied to ordinary HTE models (see ref^29^, Chapter 4), here the tests are specifically catered for our framework with instrumental variables, especially for the test based on the (modified) τ-risk.

The two tests are briefly described below. For the permutation-variance test, we aimed to compare the variance of predicted individualized treatment effect estimated from original set of covariates and permuted covariates. The rationale behind this approach is that if we permute the covariates, they should no longer to modify the treatment effects and contribute to the τ heterogeneity. In this case, if there is heterogeneity that the covariates can explain, we will have 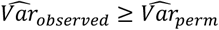 where Var denotes variance of τ. We can further set up a statistical test by repeating the permutation procedure *N* times, and the p-value can be obtained by 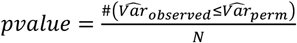 Note that we did not re-calculate the residualized Y and W; we only permute the covariates when the covariates are fit into the causal forest (or other ITE-finding procedures). The reason is that we still wish to correct *Y* and *W* for possible confounders; the permutation is employed to mimic the case in which no covariates serve as effect modifiers.

Another method we proposed is the permutation-τ-risk test, which aims to test whether introducing heterogeneity in treatment effects can lead to a better goodness-of-fit than assuming homogenous treatment effects. To assess the goodness-of-fit of a causal model, Nie et al. proposed a loss function based on Robinson transformation^9,30^, defined as,

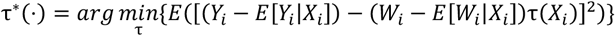

It is also referred to as the R-loss function. In the instrumental case, we propose a modification of the R-loss to

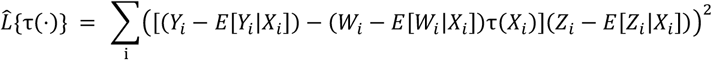

We also termed this loss as τ risk.

In addition, we define the improvement of introducing heterogeneity as,

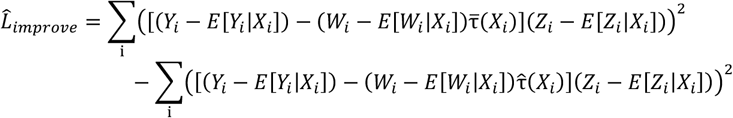

Where 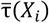 represents the average treatment effect (i.e., assume no heterogeneity in treatment effects), and 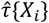 represents the individual treatment effect.

The rationale behind the permutation-τ-risk test is that we expect the total R-loss of a heterogeneous model to be *smaller* than the homogeneous model, if heterogeneity truly exists. Here we used permutation to model the null hypothesis. Intuitively, if we permute the covariates, we eliminate covariates’ contribution to the heterogeneity, which mimic the null scenario. In practice, we permute the covariates *N* iterations and recalculate the 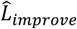. With this procedure, we can model the null model of 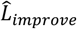 and the permutation p-value can be computed as 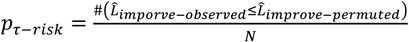. Similar to the above, the residualized Y, W and Z would not be re-calculated; only the covariates were permuted when they were fit into the forest (or other ITE-finding procedure).

### 2.8 Measuring Variable Importance

In addition to identifying the individualized treatment effect, we aimed to identify which covariate may contribute more to the heterogeneity. In other words, we wish to identify important effect modifiers that lead to differential treatment effects across different people.

We mainly used two approaches to achieve the goal. The first approach is mainly designed for the generalized random forest algorithm since it calculates the variable importance based on the split frequencies. The idea is firstly proposed by Eoghan et al.^31^, in which they define the variable importance as the frequency that a variable is selected by the causal forest algorithm to perform the split. The grf package also adopts the split-count idea and implements a simple but efficient algorithm to calculate the feature’s importance^14^, where the importance of variable *x*_*i*_ of interest up to a depth of *n* is defined as,

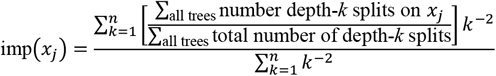

Although the split-frequency-based approach is a flexible method to identify variable importance, it has a few drawbacks. First, it can only be applied to machine learning models with node-splitting procedures, such as decision trees and random forests. Another concern is that the split-frequency approach cannot provide a reliable measurement of global feature importance owing to the inconsistency^32^. In other words, the feature’s importance calculated via ways like gain^33^ and split frequency^34^ may sometimes not be able to demonstrate the feature’s true impact on the model’s output.

Besides, considering the coming age of precision medicine, we are also interested in how much a covariate would contribute to the individual prediction, which is impossible to interpret via a split-frequency-based approach. To avoid the drawbacks of the split-frequency-based approach, we apply SHAP (<u>SH</u>apley <u>A</u>dditive ex<u>P</u>lanations) values proposed by Lundberg et al. in our framework, which can provide a robust and consistent measurement of feature importance^35^. Lundberg et al. further developed TreeExplainer to efficiently compute SHAP value for tree-based models like random forest and XGBoost^19^, which will be used by our framework to interpret the causal forest model. Briefly, the Shapley value captures each feature’s contribution after considering the rest of the features, and the Shapley value can be computed for each individual and the whole sample.

### 2.9 Quantifying effect modifiers with segmented regression

In our study, we followed a similar two-step procedure discussed by Athey et al. to explore effect modification^36^. We first visualized the relationship between the interested covariate *X*_*k*_ and estimated treatment effect τ via a scatter plot, and we performed a segmented regression to quantify the magnitude of the relationship^37^. We also fitted an ordinary least square model to test whether *X*_*k*_ was associated with τ after controlling for other covariates.

### 2.10 Individualized treatment effect estimation framework – A summary

This project aims to present a novel causal analytic framework, MR-ITE, to study the causal effect of risk factors at an individual level. The proposed framework integrates the idea of Mendelian Randomization (MR) to identify individualized treatment effects (ITEs), which reduces the risks of unmeasured confounding and reverse causality.

The MR-ITE framework comprises several main steps. First, we identify valid SNPs associated with the exposure of interest, while minimizing the risk of pleiotropic effects. Second, using the identified SNPs, we estimate a polygenic risk score (PRS) that serves as an instrumental variable. Third, we employ two approaches, including GRF and DRIV, to estimate the ITEs. These methods can potentially handle nonlinear relationships and high-dimensional data, making them suitable for estimating the ITEs in large-scale datasets. Finally, we use permutation-based methods to test for the presence of heterogeneity in the treatment effects across individuals.

### 2.11 Simulation study

We conducted two simulations to assess the performance of our proposed framework in estimating ITEs and the power of our proposed heterogeneity testing methods.

We set up a simulation study with different pleiotropic scenarios to compare our proposed framework’s performance with the regular causal forest. We also compared individualized versus constant treatment effects to demonstrate the importance of inferring individualized treatment effects when heterogeneity is present. Overall, three different pleiotropic scenarios were included in our simulations^10^:

1. Balanced pleiotropy: some genetic variants directly affect the outcome, with pleiotropic effects that are a mixture of positive and negative effects averaging to zero.
2. Directional pleiotropy: some genetic variants directly affect the outcome, with all pleiotropic effects being positive.
3. Pleiotropy via a confounder: some genetic variants affect the outcome via a confounder. In this case, the Instrument Strength Independent of Direct Effect (InSIDE) assumption is violated.

The simulation is set up following a similar idea from Burgess et al., and the data is generated as follows:

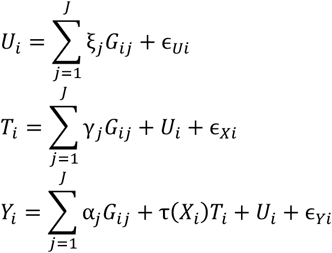

where,

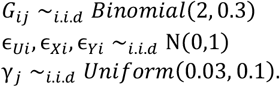

Here, *U* represents the confounders that contributes to both the treatment *T* and outcome *Y*. We simulated *X* as potential effect modifiers and it only contributes to the individualized treatment effect (τ(*X*_*i*_)). We incorporated eight different treatment effect functions τ(*X*_*i*_) to simulate the treatment effect τ; details can be found in Appendix A^38^. We simulated 100 genetic variants G_*j*_, *j* = 1, …, 100 in each scenario and considered three cases with 20, 40, and 60 invalid instruments. We simulated two types of effect modifier *X*, including continuous and binary variables. We simulated X as both continuous (standard normal) and binary (binomial with probability 0.5) variables. Following the setting from Powers et al.^38^, we simulated 50 *X*_*s*_ for scenario 1 and 2, 40 *X*_*s*_ for scenarios 3 and 4, 30 *X*_*s*_ for scenarios 5-6 and 20 *X*_*s*_ for scenarios 7-8. Among these *X*_*s*_, half of them were simulated as continuous variables, and another half as binary variables.

We set the *α*_*j*_ and ξ_*j*_ to 0 for valid instruments. For invalid instruments, *α*_*j*_ and ξ_*j*_ were set differently for different scenarios. In the balanced pleiotropy scenario (scenario 1), *α*_*j*_ was simulated from uniform(−0.1, 0.1), and the ξ_*j*_ was set to 0. For the directional pleiotropy scenario (scenario 2), *α*_*j*_ was simulated from uniform(0, 0.1), and we set ξ_*j*_ to 0. In scenario 3 (Pleiotropy via a confounder), *α*_*j*_ were set to 0 and ξ_*j*_ were drawn from uniform(−0.1, 0.1). We fit a regression forest to model the relationship between exposure and instruments, and used the prediction from the regression forest model as the instrument in the simulation^39^. The simulation was repeated 50 times in each scenario.

To avoid overfitting issues, we simulated four independent datasets with shared instruments and were used for different steps of our framework. The four datasets contained 10000 samples each. We used the first two datasets to model the association between instruments Z and outcome Y, instruments Z and treatment T, respectively, which mimic the case that we have two summary statistics results for treatment and outcome of interest. We will apply the contamination mixture approach to obtain the valid SNPs based on the first two datasets. The third dataset will be subsequently used for the training of regression forest model mentioned above, where only valid SNPs will be included in the model. The last dataset will be used as the target dataset, where we will use the causal model to predict the treatment effects.

The performance of our proposed HTE-testing methods is evaluated through simulation as well. We set up the simulation data following similar approaches discussed above, but only 40 invalid SNPs were included. Similarly, we repeated the simulation 50 times for each scenario.

To evaluate the performance of our proposed framework in capturing heterogeneity, we compared the estimated ITE with the true ITE and computed the mean squared error (MSE). Lower MSE indicates better performance. In addition, we examined the bias of the proposed approaches in estimating the ITE for the whole population and the subgroup of individuals with true ITE ranked at the top 10%.

### 2.12 Applications in real data: Heterogeneous effects of lipid traits on coronary artery disease

#### Overall analytic strategy

Using data from the UK-Biobank (UKBB) study, we applied our framework to study the heterogeneous treatment effect for several lipid-related risk factors on coronary heart disease (CAD). UK-Biobank is a large-scale cohort consisting of genetic and clinical data from ∼500,000 participants. We selected white participants with data available for principal component analysis, to minimize risks of population stratification. The current study was

#### Exposure

The main exposure is lipid levels including LDL, HDL, triglyceride and total cholesterol. They were extracted from the UKBB.

#### Outcome and covariates

CAD diagnosis was determined by International Classification of Diseases, Tenth Revision (ICD-10) code I25 in field 41202-0.0 and date in field 41262-0.0. We only considered those CAD patients with CAD diagnosis after the date of the biomarker assessment.

For covariates, we selected clinical variables likely influencing both outcomes and exposure, which can be roughly classified into three groups: biomarkers, medical history and lifestyle history (detailed in Table 2). We converted discrete variables to dummy variables, and missing data was imputed by the missRanger package^40^.

We trained two models with different covariates sets. For the first model, we only included age and gender as covariates; this mimics practical applications when there is only limited covariate information. For the second model, we additionally adjusted for multiple biomarkers and socio-demographic covariates (Table 2). With the incorporation of a larger set of covariates, we hope to identify covariates contributing to potential heterogeneity of the effect of lipids on CAD outcomes. This real-world application serves to demonstrate our framework’s ability to capture treatment effect modification/heterogeneity and inform precision prevention strategies using large-scale epidemiological data.

#### Genetic instruments

The GWAS summary statistics for lipid traits was obtained from the Global Lipids Genetics Consortium^41^. We also obtained CAD summary statistics dataset from CARDIoGRAMplusC4D Consortium^42^. In addition, we also checked that the summary statistics dataset used for PRS calculations had no overlap with the UKBB cohort.

Considering that we are using polygenic risk score as an instrument, additional quality control of genetic data is required. We followed the recommended quality control pipeline of PRSice-2 to ensure target data meets GWAS standards. Specifically, we removed SNPs with low genotyping rate (-geno 0.01), low minor allele frequency (-maf 0.001), and individuals with the low genotyping rate (-mind 0.01) ^43,44^ following the default settings. Only variants strongly associated with the exposure were included for subsequent analysis (P-value < 1e-8).

#### ITE analysis

Overall, there were 276054 subjects included in the study, 13010 of which were identified as CAD (occurring after biomarker measurement). The main outcome is the development of coronary artery disease (CAD) after the measurement of lipid levels. In our application, we also compared IV-GRF and DRIV approaches with a more standard (non-instrumental) approach, causal forest (CF, implemented in the R package GRF), in which the risk factor was directly modelled without genetic instruments.

Since so far ML-based ITE models are mainly developed for linear outcomes, we model the outcome (CAD) also as a continuous outcome, hence the treatment effects are on a linear probability scale (i.e., it reflects the changes in absolute risk or incidence of CAD per unit change of the exposure/treatment). In fact, it is not uncommon to employ linear models for binary outcomes in GWAS studies^45^, and such use may be justified by the observation that linear model is a first order Taylor approximation to a generalized linear model^46^.

As for the “treatment” variable, we considered two cases: lipid levels as a continuous and a binary treatment. In the former case of a continuous ‘treatment’, the ITE reflects the change in the absolute risk of CAD per unit increase of lipid level; whereas for a binary treatment, the ITE is the change in absolute risk of CAD for a change from dyslipidaemia (LDL>130 mg/dL^47^; TC > 220mg/dL^48^; HDL-C <46mg/dL^49^; TG>150mg/dL^47,50^) to normal levels.

### 2.13 Validation of the Heterogeneity Testing Results with Subgroup Analysis

A gold standard to validate the testing results is usually to check whether similar results can be found in an external validation dataset. However, such an external dataset is not feasible since it is difficult to find an external dataset with a comparable sample size and similar covariate sets as UKBB. Therefore, to validate our testing results, we mainly adopt the subgroup analysis approach.

Its main goal is to determine the heterogeneity of the treatment effect across subpopulations^51^. The rationale is that if heterogeneity indeed exists in the population, there must exist some subgroups whose average treatment effect is significantly different from the others. Note that the true ITE for *each individual* is not verifiable as ITE is based on a (hypothetical) counterfactual outcome; however, the treatment effects in a *subgroup* can be estimated and used to validate our ITE model.

In subgroup analysis, we first trained an ITE estimation model using a generalized random forest, and selected the ‘best representative tree’ (the tree with the lowest R-loss) as the final model to partition people into different subgroups. To retrieve clinically meaningful subgroups from the population, we further restricted the depth of the tree to 3, leading us to find four to five subgroups using the best representative tree. Using 15% of samples for training and 85% for testing, we inferred average causal effects for each subgroup via IVreg, and performed one-way ANOVA to test for differences in treatment effects across subgroups. This analysis also suggests an important clinical application of the MR-ITE framework, namely subgrouping patients/individuals with diverse responses to treatment/risk factors.

### 3.1 Simulation Study

Figure 1 illustrates the simulation outcomes for various treatment effect scenarios, considering different counts of invalid SNPs. In the balanced pleiotropy scenario, the Instrumental Causal Forest (IV-CF) significantly outperformed the regular Causal Forest (CF), as evidenced by a substantially lower Mean Squared Error (MSE) (Fig.1A). Under balanced pleiotropy, the removal of invalid pleiotropic SNPs does not appear to influence the performance of the estimator, as no significant difference is observed between methods that keep or remove the pleiotropic SNPs.

**Figure 1.**
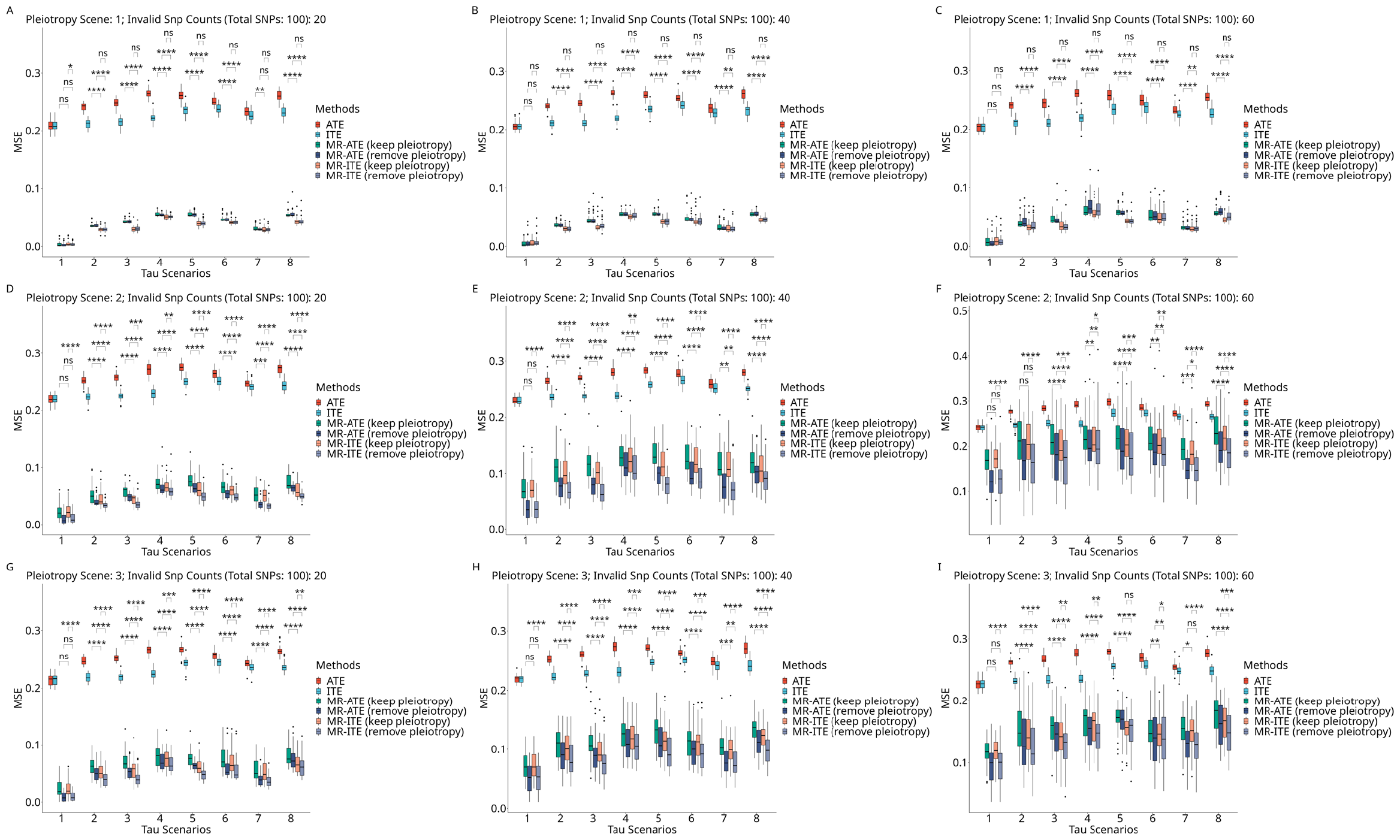
Simulation results across three different methods. Results across 8 different treatment effect scenarios, 3 pleiotropy scenarios and 3 invalid SNPs scenarios. For details of the generating distribution and scenarios, see 2.11. The 6 estimators being evaluated as follows: ATE = average treatment effect; ITE = individualized treatment effect; MR-ATE (keep pleiotropy) = MR-based average treatment effect with the presence of pleiotropy; MR-ITE (keep pleiotropy) = MR-based individualized treatment effect with the presence of pleiotropy; MR-ATE (remove pleiotropy) = MR-based average treatment effect with pleiotropy removal; MR-ITE (remove pleiotropy) = MR-based individual treatment effect with pleiotropy removal. All results presented in figure 1 are estimated using grf package. We presented paired t-test results on following comparison sets (from lowest position to the highest position): (a) MR-ATE (keep pleiotropy) vs. MR-ITE (keep pleiotropy); (b) MR-ATE (remove pleiotropy) vs. MR-ITE (remove pleiotropy); (c) MR-ITE (keep pleiotropy) vs. MR-ITE (remove pleiotropy);

We now turned to the scenarios of directional pleiotropy and pleiotropy via a confounder. As expected, IV-CF outperformed ordinary CF across all treatment effect scenarios, although the degree of improvement gradually diminished with an increase in the count of invalid SNPs. Contrary to the balanced pleiotropy scenarios, the removal of invalid pleiotropic SNPs significantly enhanced the performance of IV-CF (Fig.1D-I) under directional pleiotropy or pleiotropy via a confounder. The simulation results underscored the importance of incorporating an appropriate step for the removal of invalid pleiotropic SNPs within the framework. We further evaluated the efficacy of the ConMix approach in eliminating invalid SNPs. Our findings reveal that the ConMix approach can detect invalid SNPs with an accuracy of approximately 80% in our simulation (Supp Table 11).

To highlight the importance of inferring individualized treatment effects, we also compared the performance of the ATE and ITE estimators. ATE represents the average treatment effect which assumes a constant treatment effect across all individuals, while ITE allows the treatment effects to differ by individual. Under heterogeneous treatment effect scenarios (scenarios 2-8), the MSE results of ATE are notably higher than those of ITE (ITE vs. ATE or MR-ATE (remove pleiotropy) vs. MR-ITE (remove pleiotropy)). On the contrary, under the homogeneous treatment effect scenario (scenario 1), no significant difference is observed when ATE was compared to ITE. These findings emphasize the importance of inferring individual treatment effects in the presence of heterogeneity. To support our findings, we plotted the bias and observed that our proposed MR-ITE approach exhibits superior bias control compared to conventional methods in our simulations (Supplemental Fig.2).

In addition to benchmarking the performance of applying PRS as an instrument in inferring heterogeneous treatment effects, we conducted a simulation to validate our proposed heterogeneity-detecting methods. The results, summarized in Table 1, reveal that both methods maintain good type 1 error control in a scenario with no heterogeneity (scenario 1). They also demonstrate good power in several simple scenarios (scenarios 2, 3, 5), where the heterogeneous treatment effect functions (τ(⋅)) are simple linear combinations of the same types of covariates or exhibit weak nonlinear effects without interaction between different types of covariates. However, the permutation ττ-risk test outperforms the permutation-variance test when an interaction between different types of covariates is present (Scenarios 4, 6). Interestingly, the permutation-variance test exhibits low power in scenario 7, where a strong nonlinear effect exists in the τ(⋅), while the permutation ττ-risk maintains relatively good power in this scenario.

**Table 1:** Heterogeneity Testing Methods simulation results.

| Pleiotropy<br>Scenario | Perm-Variance Test | | | Perm-τ-risk Test | | |
| --- | --- | --- | --- | --- | --- | --- |
|  | 1 | 2 | 3 | 1 | 2 | 3 |
| Scenario 1 | 0.06 | 0.06 | 0.06 | 0 | 0.06 | 0.06 |
| Scenario 2 | 0.62 | 0.72 | 0.82 | 0.86 | 0.92 | 1 |
| Scenario 3 | 0.76 | 0.88 | 0.88 | 0.76 | 0.98 | 1 |
| Scenario 4 | 0.2 | 0.3 | 0.56 | 0.86 | 0.94 | 0.96 |
| Scenario 5 | 0.82 | 0.86 | 1 | 1 | 1 | 1 |
| Scenario 6 | 0.32 | 0.34 | 0.54 | 0.88 | 0.86 | 0.92 |
| Scenario 7 | 0.1 | 0.04 | 0.14 | 0.52 | 0.52 | 0.7 |
| Scenario 8 | 0.5 | 0.46 | 0.76 | 1 | 1 | 1 |

The table shows the simulation results of two permutation-based heterogeneity testing methods, the permutation-variance test, and the permutation-τ-risk test. Scenario 1 is a scenario with homogeneous treatment effect, and the results refer to the type I error of the method. The rest of the scenarios show heterogeneous treatment effect, and the power will be used to denote the performance of the test in detecting heterogeneity.

**Table 2:** Covariates included in the model 2 analysis.

| Traits | Shared Discrete Variables | Shared Continuous Variables | Lipid-related variables |
| --- | --- | --- | --- |
| LDL-C | Gender,<br>Non-alcohol drinker,<br>Previous alcohol drinker,<br>Current alcohol drinker,<br>Non-smoker,<br>Previous smoker,<br>Current smoker,<br>Blood pressure medication history,<br>Cholesterol lowering medication,<br>Hypertension history,<br>Type 2 diabetes history,<br>Heart failure history,<br>Hemorrhage Stroke history,<br>Ischemic Stroke history | Principal Component 1-10,<br>Townsend Deprivation Index,<br>diastolic blood pressure,<br>systolic blood pressure,<br>BMI, Weight, Body-fat-percentage, Age,<br>Creatinine,<br>C-reactive protein,<br>Cystatin C,<br>Gamma glutamyl transferase,<br>Aspartate aminotransferase,<br>Glucose, HbA1c, Calcium<br>Direct bilirubin, Urea, IGF-1, Phosphate,<br>SHBG, Testosterone, Urate, Vitamin D,<br>Total bilirubin, Total protein | HDL-C,<br>Apolipoprotein A,<br>Triglycerides |
| HDL-C |  |  | LDL-C,<br>Apolipoprotein B,<br>Lipoprotein A,<br>Triglycerides, |
| Total Cholesterol |  |  | Triglycerides |
| Triglyceride |  |  | NA |

The table summarizes the covariates used in different traits models. We cluster the covariates into three groups, discrete variables, continuous variables and lipid-related variables. To avoid potential pleiotropy via confounder issue, we specify lipid-related covariates for each traits separately, where the corresponding lipoprotein will be excluded.

### 3.2 Treatment effect of lipid-related traits on coronary artery disease

#### 3.2.1 Baseline Characteristics of included participants

The baseline characteristics of the study’s continuous and categorical covariates are summarized in the supplementary materials. A partial F-test was conducted to evaluate the strength of the polygenic risk score as an instrument^52^. The results indicate that the polygenic risk score can be considered a strong instrument, as the F-statistics in all cases significantly exceed 10 (refer to Table 3). The covariates included for various lipid-related traits are also outlined in the supplementary materials, with most covariates being common across different traits. We also compared the estimate of overall treatment effect based on standard regression against that from an ordinary regression, using the Wu-Hausman test as implemented in IVreg^53,54,55^. If the null hypothesis is rejected, it indicates that the explanatory variable is endogenous. In this case, the IV estimator is consistent, while the standard regression estimator is not. Conversely, if the null hypothesis is not rejected, the IV and the ordinary regression estimator are considered to be both consistent, although the IV estimator has a larger variance. The original regression estimator is preferred in this case. Our results suggest that the IV estimator is preferred in the studies of LDL-C and Total Cholesterol (refer to Table 3). Consequently, our discussion primarily focuses on the results of these studies.

**Table 3:** Partial F-test and Wu-Hausman Test to Assess Instrument Strength and the difference between MR and observational Estimates.

|  |  | Weak Instrument Test |  | Wu-Hausman Test |  |
| --- | --- | --- | --- | --- | --- |
|  |  | Statistics | P-value | Statistics | P-value |
| Model 1 | LDL-C | 10302.84 | < 2e-16 | 32.66 | **<u>1.1e-08</u>** |
|  | HDL-C | 3969.22 | < 2e-16 | 0.11 | 0.741 |
|  | Total Cholesterol | 7480.83 | < 2e-16 | 26.44 | **<u>2.72e-07</u>** |
|  | Triglyceride | 6346.991 | < 2e-16 | 4.676 | **<u>0.0306</u>** |
| Model 2 | LDL-C | 17326.107 | < 2e-16 | 6.746 | **<u>0.0094</u>** |
|  | HDL-C | 3331.451 | < 2e-16 | 0.049 | 0.824 |
|  | Total Cholesterol | 10818.20 | < 2e-16 | 7.09 | **<u>0.00775</u>** |
|  | Triglyceride | 1747.035 | < 2e-16 | 1.199 | 0.273 |

The table shows the partial F-test results for assessing instrument strength and Wu-Hausman test for assessing whether the effect estimates from the instrumented approach (MR) are significantly different from those under an observational approach. We assess the strength of instrument in multiple lipid-traits to disease setting.

#### 3.2.2 LDL-C

##### 3.2.2.1 LDL-C Imposes Heterogeneous Effects on CAD

We initially utilized our framework to investigate the causal association between LDL-C and CAD under both continuous and binary exposure scenarios. The findings from our study reaffirmed that an increase in LDL-C levels is causally linked to an increased risk of CAD. This association was consistently observed across two distinct models (refer to Fig. 2A, 2B, and Supplementary Feig. 5A, 5B).

**Figure 2.**
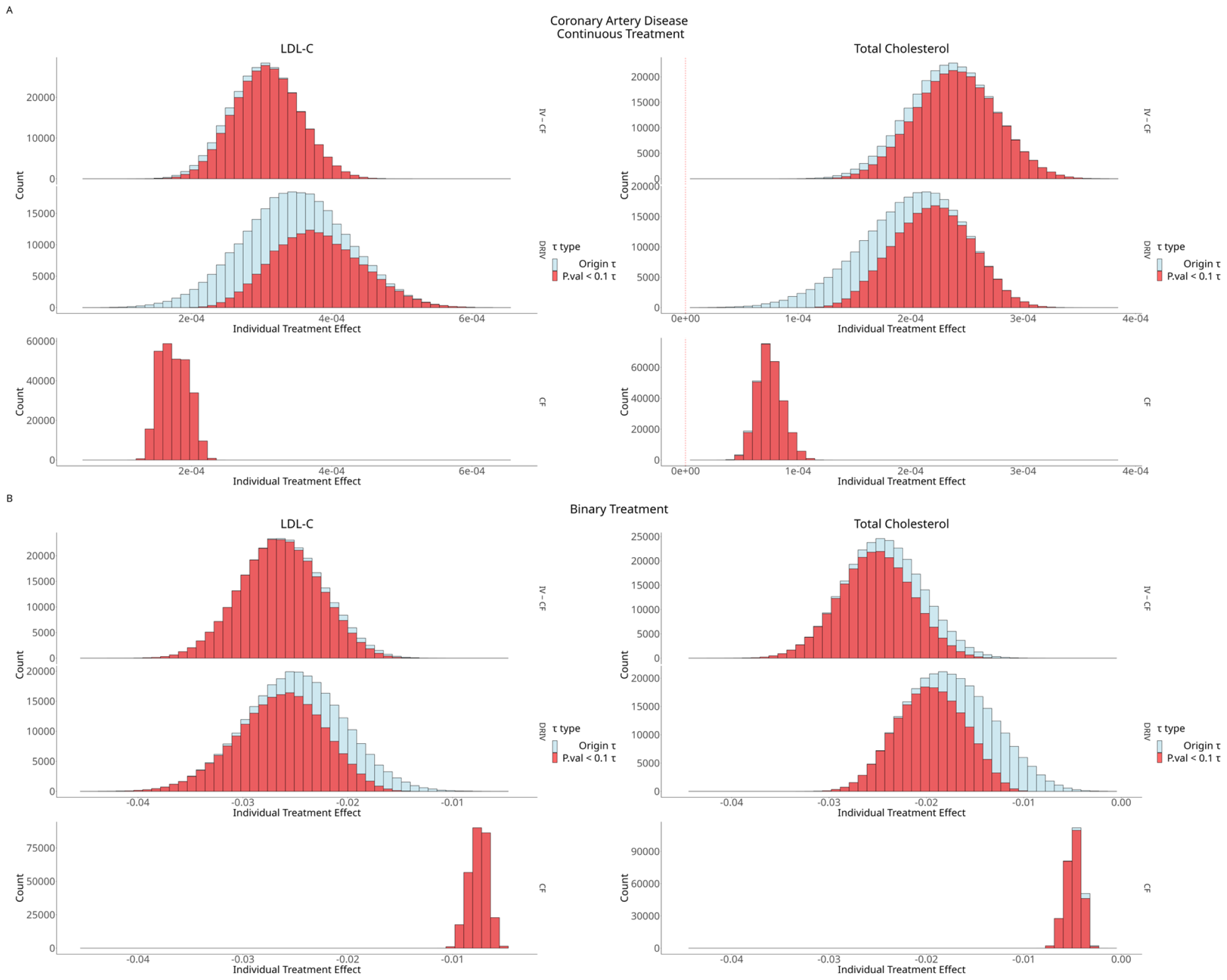
Predicted Treatment Effect of LDL-C and Total Cholesterol on CAD (Model 2). Histogram results of applying DRIV, IVCF and CF in estimating the individualized causal effect of LDL-C/Total Cholesterol on CAD incorporating covariate set 3. A: Estimating the individualized causal effect with a continuous treatment setting. B: Estimating the individualized causal effect with a binary treatment setting.

In our study on CAD, we observed that IV-GRF, DRIV, and Causal Forest predict positive treatment effects for all participants. Notably, IV-GRF and DRIV yielded higher predictions than CF in Model 2. Additionally, DRIV detected a less significant treatment effect compared to IV-GRF. In Model 1, the CF approach failed to detect a significant treatment effect in most patients, with individualized treatment effects centered around zero. We hypothesize that this is due to the inclusion of only two covariates, age and gender, in Model 1, leading to a failure in controlling for potential confounders. This is contrary to the mainstream finding that an increment of LDL-C imposes a higher risk of CAD. Our results support the assertion that incorporating an instrument in an observational study can enhance the accuracy of Individualized Treatment Effect (ITE) estimation. It’s important to note that we modeled the original risk factor without using instruments under the CF approach, as previously described. In Model 2, analyses using IV-GRF and DRIV indicate that a per unit (1 mg/dL) increment of LDL-C increases the individual CAD risk by approximately 0.03%. In contrast, for CF, the average risk increase was less than 0.02% (Fig. 2A LDL-C). Under a ‘binary treatment’ scenario (normal lipid levels vs. hyperlipidemia), IV-GRF/DRIV also yielded higher treatment effect predictions. Our findings suggest that reducing LDL-C levels to a normal range below the optimal cutoff (130 mg/dL) could lead to a roughly 3% reduction in CAD incidence.

Intriguingly, we found that the Causal Forest (CF) only detects an average CAD risk reduction of approximately 0.5% in the ‘binary treatment’ setting. This is significantly less than what is detected in the MR-ITE framework (Fig. 2B). This pattern aligns with our expectations, as the results of the Wu-Hausman test indicate that only the IV estimator can return a consistent estimation. Our findings are corroborated by several studies. For instance, Brian et al. reported an absolute risk reduction in Atherosclerotic Cardiovascular Disease (ASCVD) ranging from 2.1% to 8.6% for patients whose LDL-C levels were controlled under 100 mg/dL following LDL-C reduction therapy^56^. Notably, compared to ordinary MR methods, our proposed MR-ITE framework enables the estimation of a causal effect for *each individual*.

We further evaluated the heterogeneity of treatment effects using our proposed permutation-based tests. Our findings indicate that LDL-C modification results in heterogeneous effects on CAD in both continuous and binary treatment settings (refer to Table 4). These findings are supported by several Randomized Controlled Trial (RCT) studies. For instance, Pravastatin was reported to exhibit heterogeneous efficacy in reducing coronary events^57^. It was found that the effect of Pravastatin in reducing coronary events varies between females and males. Moreover, patients with higher pretreatment levels of LDL-C were observed to receive a greater protective effect from Pravastatin. This phenomenon may corroborate our finding that LDL-C manipulation results in a heterogeneous effect on CAD, with the heterogeneity of the treatment effect potentially attributable to several covariates, such as gender. Another noteworthy finding is from a study by Oscar et al., who identified heterogeneity in cost-effectiveness ratios after adjusting for absolute risk^58^. This underscores the importance of a personalized statin prescription policy for CAD prevention in the population.

**Table 4:** Permutation-based test to assess the presence of heterogeneity.

|  |  | Coronary Artery Disease |  |
| --- | --- | --- | --- |
|  |  | Perm-Var test | Perm-Risk test |
| Continuous Trait | LDL-C | 0.01 | 0.01 |
|  | Total Cholesterol | 0.01 | 0 |
| Binary Trait | LDL-C | 0.02 | 0 |
|  | Total Cholesterol | 0 | 0.01 |

The table summarizes two permutation-based test results for assessing the presence of heterogeneity in different lipid-trait to disease setting.

**Table 5:** Model 2 Tau Summary (Continuous Trait)

|  |  | Positive<br>Tau | Significant<br>Tau<br>(P-value <<br>0.1) | Min | 5% | 10% | 25% | Median | 75% | 90% | 95% | Max | 50%<br>Coverage<br>(25% -<br>75%) | 80%<br>Coverage<br>(10% -<br>90%) | 90%<br>Coverage<br>(5% - 95%) | Mean |
| --- | --- | --- | --- | --- | --- | --- | --- | --- | --- | --- | --- | --- | --- | --- | --- | --- |
| LDL-C | CF | 100.00% | 100.00% | 1.190E-03 | 1.454E-03 | 1.497E-03 | 1.586E-03 | 1.737E-03 | 1.909E-03 | 2.024E-03 | 2.081E-03 | 2.419E-03 | 3.226E-04 | 5.267E-04 | 6.275E-04 | 1.750E-03 |
|  | DRIV | 100.00% | 55.94% | 3.867E-04 | 2.241E-03 | 2.515E-03 | 2.977E-03 | 3.483E-03 | 3.990E-03 | 4.458E-03 | 4.728E-03 | 6.622E-03 | 1.013E-03 | 1.943E-03 | 2.486E-03 | 3.484E-03 |
|  | IVCF | 100.00% | 94.86% | 1.118E-03 | 2.255E-03 | 2.424E-03 | 2.715E-03 | 3.047E-03 | 3.378E-03 | 3.672E-03 | 3.845E-03 | 5.060E-03 | 6.634E-04 | 1.248E-03 | 1.590E-03 | 3.047E-03 |
| HDL-C | CF | 100.00% | 99.98% | -4.262E-03 | -3.471E-03 | -3.316E-03 | -3.017E-03 | -2.628E-03 | -2.189E-03 | -1.818E-03 | -1.646E-03 | -9.956E-04 | 8.277E-04 | 1.498E-03 | 1.825E-03 | -2.598E-03 |
|  | DRIV | 99.96% | 14.03% | -7.429E-03 | -4.795E-03 | -4.367E-03 | -3.617E-03 | -2.803E-03 | -2.078E-03 | -1.515E-03 | -1.217E-03 | 6.381E-04 | 1.539E-03 | 2.852E-03 | 3.579E-03 | -2.881E-03 |
|  | IVCF | 97.34% | 3.69% | -1.227E-02 | -6.854E-03 | -6.125E-03 | -4.938E-03 | -3.643E-03 | -2.368E-03 | -1.226E-03 | -5.452E-04 | 5.590E-03 | 2.570E-03 | 4.899E-03 | 6.309E-03 | -3.661E-03 |
| Total<br>Cholesterol | CF | 100.00% | 99.11% | 3.167E-04 | 5.698E-04 | 6.060E-04 | 6.681E-04 | 7.415E-04 | 8.233E-04 | 9.032E-04 | 9.508E-04 | 1.280E-03 | 1.552E-04 | 2.973E-04 | 3.810E-04 | 7.486E-04 |
|  | DRIV | 100.00% | 69.62% | -2.107E-05 | 1.210E-03 | 1.403E-03 | 1.717E-03 | 2.045E-03 | 2.338E-03 | 2.572E-03 | 2.697E-03 | 3.456E-03 | 6.207E-04 | 1.168E-03 | 1.487E-03 | 2.012E-03 |
|  | IVCF | 100.00% | 89.78% | 8.537E-04 | 1.735E-03 | 1.872E-03 | 2.104E-03 | 2.363E-03 | 2.621E-03 | 2.845E-03 | 2.974E-03 | 3.863E-03 | 5.168E-04 | 9.728E-04 | 1.239E-03 | 2.360E-03 |
| Triglycerid<br>e | CF | 100.00% | 80.98% | 5.382E-05 | 1.734E-04 | 1.897E-04 | 2.193E-04 | 2.553E-04 | 2.915E-04 | 3.226E-04 | 3.407E-04 | 4.666E-04 | 7.224E-05 | 1.329E-04 | 1.672E-04 | 2.558E-04 |
|  | DRIV | 75.86% | 0.00% | -1.593E-03 | -3.960E-04 | -2.401E-04 | 1.117E-05 | 2.826E-04 | 5.489E-04 | 7.895E-04 | 9.342E-04 | 2.224E-03 | 5.377E-04 | 1.030E-03 | 1.330E-03 | 2.775E-04 |
|  | IVCF | 21.99% | 0.58% | -1.590E-03 | -7.762E-04 | -6.616E-04 | -4.677E-04 | -2.502E-04 | -3.096E-05 | 1.658E-04 | 2.835E-04 | 1.097E-03 | 4.367E-04 | 8.274E-04 | 1.060E-03 | -2.490E-04 |

The table summarizes the estimated individualized treatment effects for different lipid-trait to disease models under continuous treatment setting. Tau indicates the increase in probability of outcome for every one-unit (10mg/dL) increase of the lipid level.

**Table 6:** Model 2 Tau Summary (Binary Trait)

|  |  | Negative<br>Tau | Significant<br>Tau<br>(P-value <<br>0.1) | Min | 5% | 10% | 25% | Median | 75% | 90% | 95% | Max | 50%<br>Coverage<br>(25% -<br>75%) | 80%<br>Coverage<br>(10% -<br>90%) | 90%<br>Coverage<br>(5% - 95%) | Mean |
| --- | --- | --- | --- | --- | --- | --- | --- | --- | --- | --- | --- | --- | --- | --- | --- | --- |
| LDL-C | CF | 100.00% | 100.00% | -1.076E-02 | -9.056E-03 | -8.748E-03 | -8.156E-03 | -7.464E-03 | -6.879E-03 | -6.430E-03 | -6.173E-03 | -3.996E-03 | 1.277E-03 | 2.318E-03 | 2.882E-03 | -7.529E-03 |
|  | DRIV | 100.00% | 73.81% | -4.568E-02 | -3.304E-02 | -3.126E-02 | -2.829E-02 | -2.508E-02 | -2.192E-02 | -1.910E-02 | -1.743E-02 | -4.850E-03 | 6.372E-03 | 1.215E-02 | 1.561E-02 | -2.513E-02 |
|  | IVCF | 100.00% | 96.63% | -4.283E-02 | -3.265E-02 | -3.124E-02 | -2.891E-02 | -2.627E-02 | -2.358E-02 | -2.117E-02 | -1.979E-02 | -1.082E-02 | 5.334E-03 | 1.007E-02 | 1.285E-02 | -2.625E-02 |
| HDL-C | CF | 0.00% | 100.00% | 5.405E-03 | 6.774E-03 | 7.040E-03 | 7.568E-03 | 8.186E-03 | 8.759E-03 | 9.232E-03 | 9.495E-03 | 1.120E-02 | 1.191E-03 | 2.191E-03 | 2.721E-03 | 8.162E-03 |
|  | DRIV | 13.16% | 0.88% | -1.595E-02 | -2.571E-03 | -8.170E-04 | 2.292E-03 | 6.029E-03 | 1.000E-02 | 1.363E-02 | 1.572E-02 | 2.929E-02 | 7.709E-03 | 1.444E-02 | 1.829E-02 | 6.226E-03 |
|  | IVCF | 10.19% | 0.86% | -2.978E-02 | -3.843E-03 | -1.080E-04 | 6.368E-03 | 1.400E-02 | 2.228E-02 | 3.018E-02 | 3.512E-02 | 7.274E-02 | 1.591E-02 | 3.029E-02 | 3.897E-02 | 1.462E-02 |
| Total<br>Cholesterol | CF | 100.00% | 97.14% | -8.263E-03 | -6.291E-03 | -5.992E-03 | -5.439E-03 | -4.809E-03 | -4.259E-03 | -3.849E-03 | -3.628E-03 | -2.170E-03 | 1.180E-03 | 2.142E-03 | 2.664E-03 | -4.869E-03 |
|  | DRIV | 100.00% | 68.25% | -3.338E-02 | -2.473E-02 | -2.339E-02 | -2.091E-02 | -1.783E-02 | -1.455E-02 | -1.167E-02 | -1.003E-02 | -1.001E-04 | 6.366E-03 | 1.172E-02 | 1.470E-02 | -1.766E-02 |
|  | IVCF | 100.00% | 82.87% | -4.498E-02 | -3.111E-02 | -2.968E-02 | -2.726E-02 | -2.453E-02 | -2.177E-02 | -1.931E-02 | -1.783E-02 | -8.020E-03 | 5.489E-03 | 1.037E-02 | 1.328E-02 | -2.451E-02 |
| Triglycerid<br>e | CF | 100.00% | 70.43% | -6.121E-03 | -4.517E-03 | -4.279E-03 | -3.865E-03 | -3.386E-03 | -2.883E-03 | -2.421E-03 | -2.148E-03 | -2.828E-04 | 9.828E-04 | 1.858E-03 | 2.369E-03 | -3.365E-03 |
|  | DRIV | 40.30% | 0.87% | -2.541E-02 | -8.080E-03 | -5.941E-03 | -2.438E-03 | 1.408E-03 | 5.478E-03 | 9.837E-03 | 1.298E-02 | 3.648E-02 | 7.916E-03 | 1.578E-02 | 2.106E-02 | 1.750E-03 |
|  | IVCF | 11.76% | 0.38% | -1.815E-02 | -2.860E-03 | -5.994E-04 | 3.219E-03 | 7.529E-03 | 1.183E-02 | 1.562E-02 | 1.780E-02 | 3.223E-02 | 8.609E-03 | 1.622E-02 | 2.066E-02 | 7.513E-03 |

The table summarizes the estimated individualized treatment effects for different lipid-trait to disease models under binary treatment setting. Tau indicates the increase in probability of outcome for controlling the lipid level under the optimal threshold, in other words, we compare a favourable lipid profile (coded as 1) vs an unfavorable lipid profile (coded as 0), analogous to having received a treatment to improve dyslipidaemia. Therefore, the direction of tau is reversed compared to the continuous treatment setting.

Additionally, our findings of heterogeneity are supported by the subgroup analysis. We observed a significant ANOVA p-value in both binary and continuous treatment settings (Continuous Model: ANOVA p-value = 0.0241; Binary Model: ANOVA p-value = 0.0273) (refer to Supplementary Taables 4 and 6). This suggests significantly different treatment across the subgroups identified by our MR-ITE framework.

##### 3.2.2.2 Features that contribute to the heterogeneity of LDL-C on CAD

Beyond the identification of heterogeneity in the impact of LDL-C on CAD, our interest extends to the covariates that contribute to this heterogeneity. Figures 3A and 3C depict the SHAP patterns of the top 10 most important clinical features, as identified through the DRIV model of LDL-C’s influence on CAD. These patterns are presented under both continuous and binary treatment settings, utilizing a beeswarm plot for visualization. Furthermore, we segmented the population into deciles based on the corresponding feature values, enabling the visualization of potential effect modifiers.

**Fig 3.**
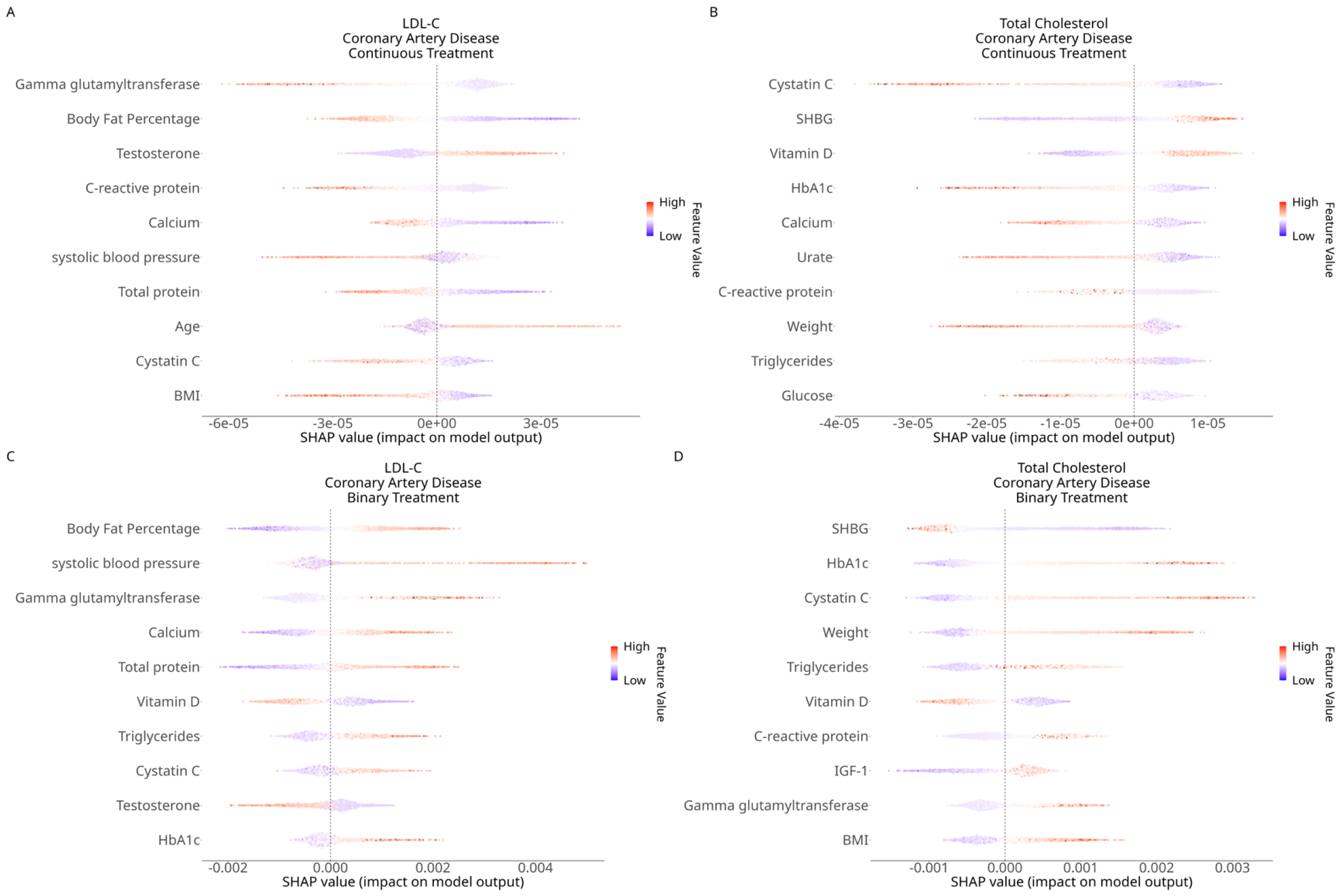
Overall top 10 clinical effect modifier on CAD identified by LDL-C/TC’s causal model with SHAP analysis. Beeswarmplot of top 10 important covariates identified under different scenarios with SHAP analysis. A: LDL-C, continuous treatment scenario. B: Total Cholesterol, continuous treatment scenario. C: LDL-C, binary treatment scenario. D: Total Cholesterol, binary treatment scenario.

Primarily, we discovered that the body fat percentage is the most significant variable in the model under both continuous and binary treatment settings (Fig.3A, C). We observed that LDL-C reduction in patients with a higher body fat percentage showed weaker protective effect on CAD, as opposed to those with lower fat percentage. This pattern is also evident in other obesity indicators such as BMI (Fig.4E, 4F, 4S, 4T). These findings align with several studies that highlight the robust relationship between obesity and CAD. For example, Sandfort et al. demonstrated that obese patients with hyperlipidemia experience more severe atheroma progression despite optimized statin therapy^59^. The authors suggested that hyperlipidemic patients with obesity may have an elevated risk of CAD, as atheroma progression can lead to atherosclerosis, the primary cause of CAD^60^. This also elucidates our observation that obese patients derive less benefit from LDL-C reduction. Our results strongly advocate for a combination therapy of weight and LDL-C reduction to achieve a more substantial protective effect against CAD.

**Fig 4.**
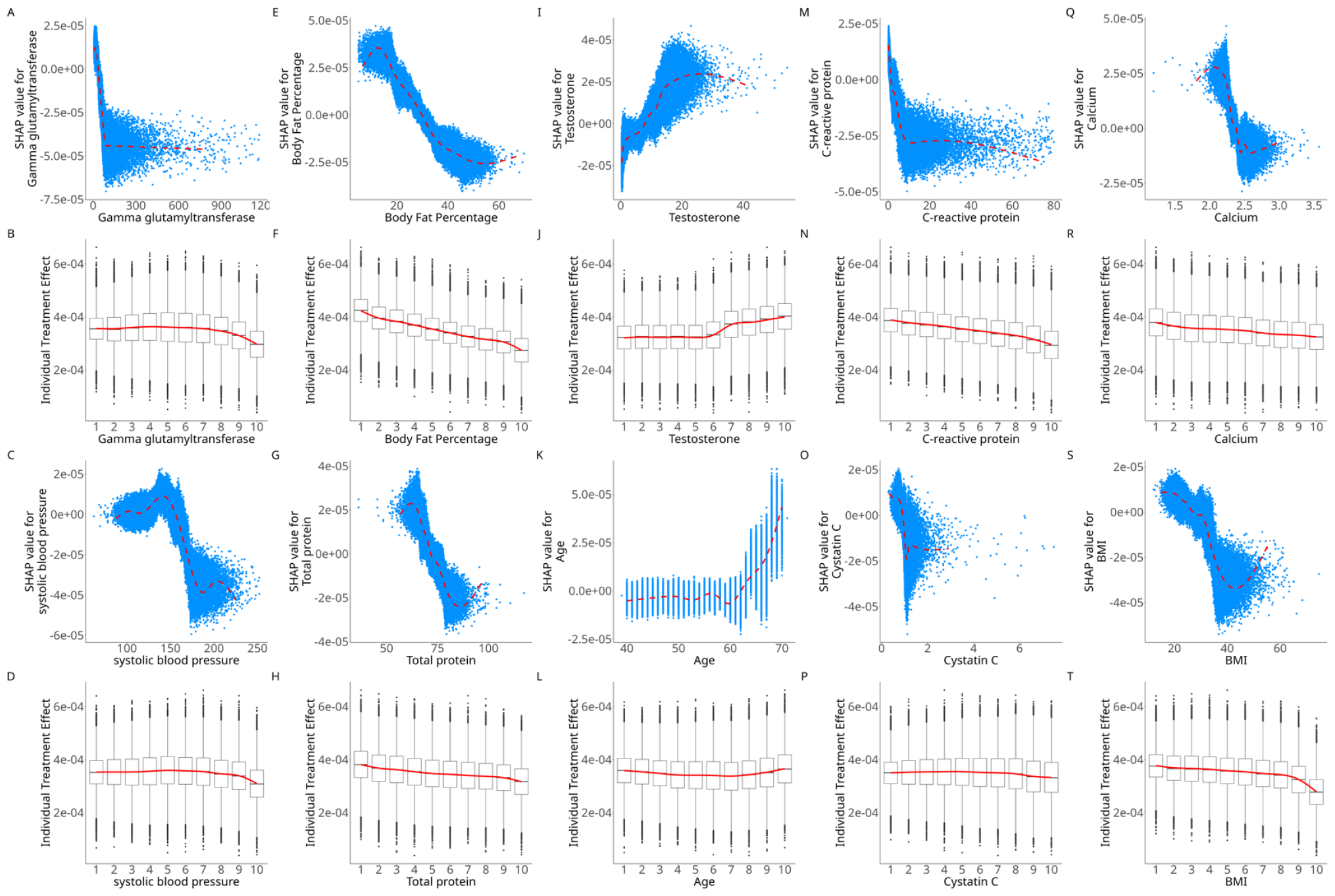
Shapley value plot with LDL-C as risk factor on CAD in continuous treatment setting. A, E, I, M, Q, C, G, K, O, S: Scatterplots of SHAP value (y-axis) versus observed value (x-axis) of top 10 important covariates identified under continuous treatment scenario where LDL-C as risk factor and CAD as the outcome of interest. B, F, J, N, R, D, H, L, P, T: Boxplots of estimated individual treatment effects (y-axis) versus observed value (x-axis) of top 10 important covariates identified under continuous treatment scenario where LDL-C as risk factor and CAD as the outcome of interest.

Furthermore, we identify systolic blood pressure as an important variable, akin to the obesity-related covariates previously mentioned, in both continuous and binary treatment settings (Fig.3A, 3C, 4C, 4D, 5E, 5F). Numerous studies have established hypertension as one of the most potent risk factors for cardiovascular diseases, including coronary artery disease^61,62^. Our research suggests that hypertension may act as an effect modifier of LDL-C’s impact on CAD. We note a reduced protective treatment effect in the population with systolic blood pressure in the top 10% (Fig.5F), and the SHAP analysis yields a positive SHAP estimation for systolic blood pressure exceeding 150 in the binary model. The segmented least square analysis further corroborates that the optimal threshold is approximately 150. These findings imply that hypertension could significantly weaken the protective effect of lowering LDL-C against CAD. Our results also endorse the assertion that a combination therapy of LDL-C and blood pressure-lowering agents is associated with a lower risk of CAD compared to mono-therapy^63^.

**Fig 5.**
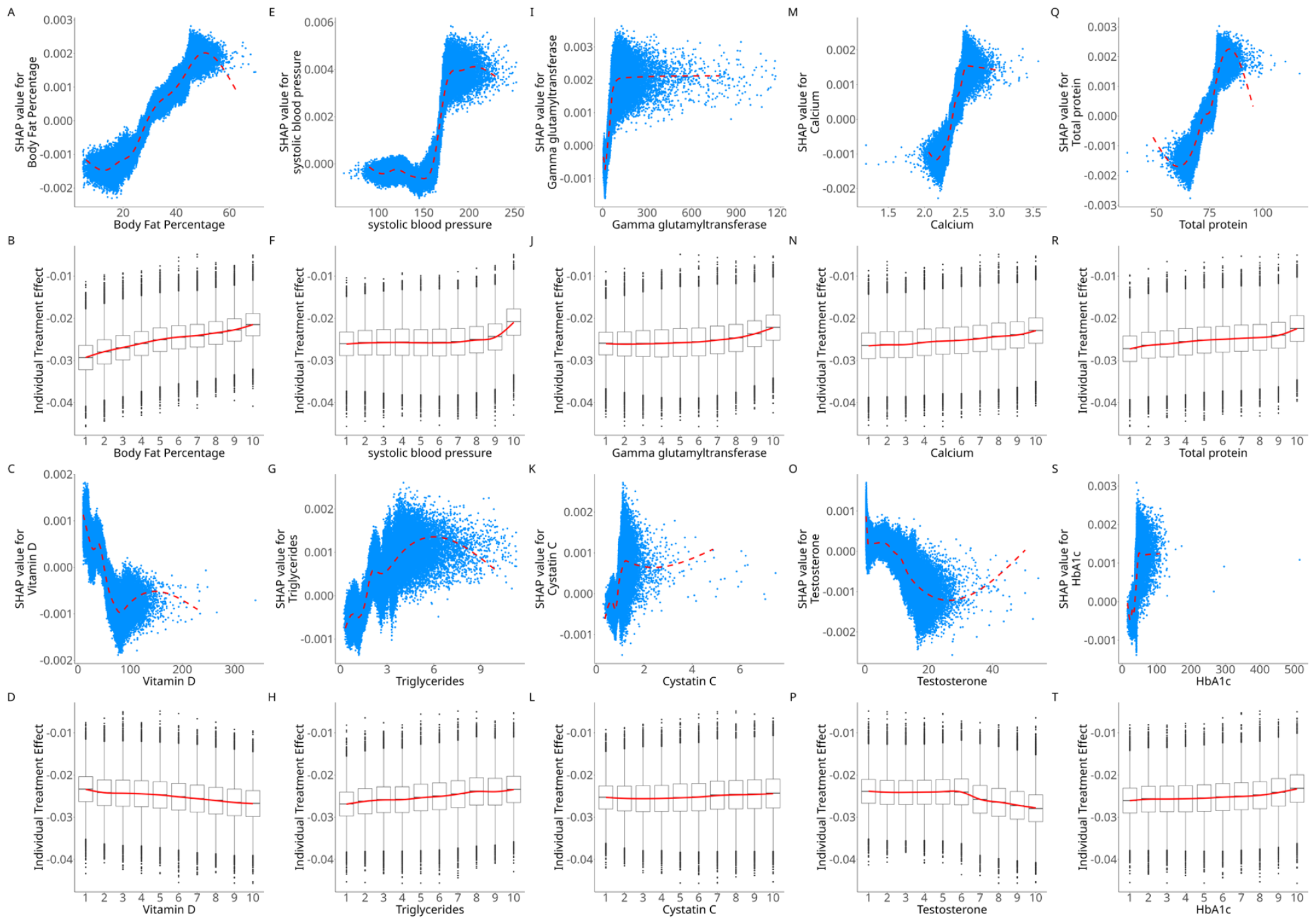
Shapley value plot with LDL-C as risk factor on CAD in binary treatment setting. A, E, I, M, Q, C, G, K, O, S: Scatterplots of SHAP value (y-axis) versus observed value (x-axis) of top 10 important covariates identified under binary treatment scenario where LDL-C as risk factor and CAD as the outcome of interest. B, F, J, N, R, D, H, L, P, T: Boxplots of estimated individual treatment effects (y-axis) versus observed value (x-axis) of top 10 important covariates identified under binary treatment scenario where LDL-C as risk factor and CAD as the outcome of interest.

Furthermore, we identified age and testosterone as significant variables in the CAD DRIV model under both continuous treatment settings (Fig.3A). We observed that the SHAP value gradually increases after 60 years of age, and the segmented least square analysis also indicated that age becomes a stronger effect modifier after the threshold of 60 years (Fig.4K, 4L). This observation is supported by other studies. For instance, Nozue et al. compared coronary atherosclerosis and vascular responses to statin therapy between elderly (> 65 years old) and non-elderly individuals^64^. They found that coronary atherosclerosis was more advanced in elderly patients, which aligns with our finding that elderly patients exhibit a higher CAD risk in response to an increase in LDL-C. LDL-C is considered to be independently associated with the presence and extent of early systemic atherosclerosis in the absence of major cardiovascular risk factors (CVRFs)^65^.

Testosterone exhibits a similar pattern to age (Fig.4I, 4J), suggesting that males may receive a larger protective effect against CAD with the decrease of LDL-C to a normal range. This finding is consistent with a study showing that statin treatment is less effective in improving the plasma lipid profile in dyslipidemic women compared to men^66^, which may leave impact on clinical outcomes. Moreover, Petretta et al. demonstrated that statin therapy can reduce the risk of CHD events in men without prior cardiovascular disease, while its effect on women is less significant^67^. This is consistent with our finding regarding testosterone’s pattern in modifying the treatment effect of LDL-C on CAD.

Another finding is that the serum calcium level is identified as the primary variable contributing to the heterogeneity of LDL-C’s causal effect on CAD in both continuous and binary models. Our results indicate that a high level of calcium corresponds to a positive SHAP value under the binary model (Fig.4Q, 4R, 5M, 5N), suggesting an inverse relationship with the protective effect of lowering LDL-C against CAD. This aligns with several studies that have highlighted the association between genetically elevated serum calcium and increased odds of coronary artery disease and myocardial infarction^68,69^. Interestingly, our results also suggest that Vitamin D, which exhibits a pattern inverse to that of serum calcium, is another significant variable. Numerous studies have reported a positive correlation between Vitamin D deficiency and the pathogenesis of CAD^70,71^. We found that an elevated Vitamin D level corresponds to a negative SHAP under the binary treatment analysis, implying that it can enhance the protective effect of lowering LDL-C against CAD. Further studies have also shown that an increased level of Vitamin D intervention is associated with lower systolic blood pressure^72^, supporting our finding regarding the association between the treatment effect of lowering LDL-C and the levels of Vitamin D and systolic blood pressure.

Also, we observed that Cystatin C, a marker of renal dysfunction, is among the top effect modifiers in both the continuous and binary LDL-C models (Fig.4O, 4P, 5K, 5L). Existing research already demonstrates that elevated serum Cystatin C levels are associated with an increased burden of coronary atherosclerotic plaque, indicating its causal effect on the heightened risk of coronary atherosclerosis^73^. Our findings reveal that a higher Cystatin C level correlates with a diminished protective treatment effect (Fig.5K, 5L). These results are supported by other studies. For example, Pontremoli et al. highlighted that reducing LDL-C in patients with Chronic Kidney Disease (CKD) can be beneficial in preventing major atherosclerotic events^74^.

#### 3.2.3 Total Cholesterol

##### 3.2.3.1 Total Cholesterol Increment Heterogeneously Increases the Risk of CAD

In addition to LDL-C, we also investigated whether total cholesterol is a causal risk factor for CAD. Elevated cholesterol levels in the blood can precipitate atherosclerosis, potentially increasing the risk of CAD^75^. Our ITE analysis reveals an increased risk of CAD associated with each unit increment of total cholesterol (Fig.2A Total Cholesterol, Supplemental Fig.5A Total Cholesterol).

In parallel with LDL-C, the causal forest framework in Model 1 did not detect a significant treatment effect. This observation supports our assertion that the inclusion of an instrument could enhance causal effect estimation in the presence of unobserved confounders. Our findings align with several meta-analyses and prospective studies^75,76^. Furthermore, we also observe a similar risk increment between altering LDL-C and total cholesterol. This observation might suggest a similar underlying mechanism influencing these particles’ effect on CAD. We also evaluated heterogeneity using our proposed test and detected heterogeneity in individualized treatment effects. This finding indicates that the effects of total cholesterol on CAD vary across the population. Our subgroup analysis further corroborates these findings, as the ANOVA p-values for both models are less than 0.05 (Supplemental Tables 8 and 10). This evidence suggests that the effects of total cholesterol on CAD are indeed heterogeneous across different subgroups within the population.

##### 3.2.3.2 Features that Contribute to the Individualized Treatment Effect of Total Cholesterol on CAD

In our analysis, we utilized SHAP to evaluate the top variables (Fig.3B, 3D). Notably, there was a significant overlap among the top 10 variables between the LDL-C and TC models in both continuous and binary treatment settings. For example, BMI and weight emerged as key variables in the LDL-C model, mirroring the pattern observed for body fat percentage. This suggests that obesity is a critical variable in the total cholesterol model as well. Furthermore, Vitamin D and Calcium exhibited a similar pattern in the total cholesterol model as seen in the LDL-C model. The substantial overlap of key variables between the LDL-C and TC models suggests a common underlying mechanism influencing CAD risk, potentially linked to progressive atherogenesis under elevated LDL-C/TC levels.

A noteworthy observation from our study is that C-reactive protein (CRP) emerged as the most important variable in the total cholesterol model. Numerous studies have identified CRP as a crucial predictor of future CAD^77-79^. The association between elevated CRP levels and increased CAD risk may be attributed to arterial inflammation. CRP binds to Low-Density Lipoprotein (LDL) and is present in atherosclerotic plaques, potentially contributing to CAD onset^80^. Our findings suggest that individuals with higher CRP levels may derive less benefit from the protective effect against CAD offered by lowering total cholesterol. Interestingly, Insulin-like Growth Factor 1 (IGF-1) was also identified as a key variable in the total cholesterol model, exhibiting a pattern similar to CRP. This is corroborated by a Mendelian randomization study which found that elevated serum IGF-1 levels are associated with a higher CAD risk^81^.

Surprisingly, our study identified Sex Hormone-Binding Globulin (SHBG) as the most top variable in both the binary and continuous TC-CAD models, ranking 1st and 2nd respectively. Previous research has indicated that participants with a high number of cardiovascular risk factors tend to exhibit lower SHBG levels^82^. Furthermore, an inverse correlation has been observed between SHBG and high C-reactive protein levels^83^, a finding that aligns with the pattern we observed between C-reactive protein and SHBG (Fig.6G, 6H, 6E, 6F, 7A, 7B, 7G, 7H).

**Fig 6.**
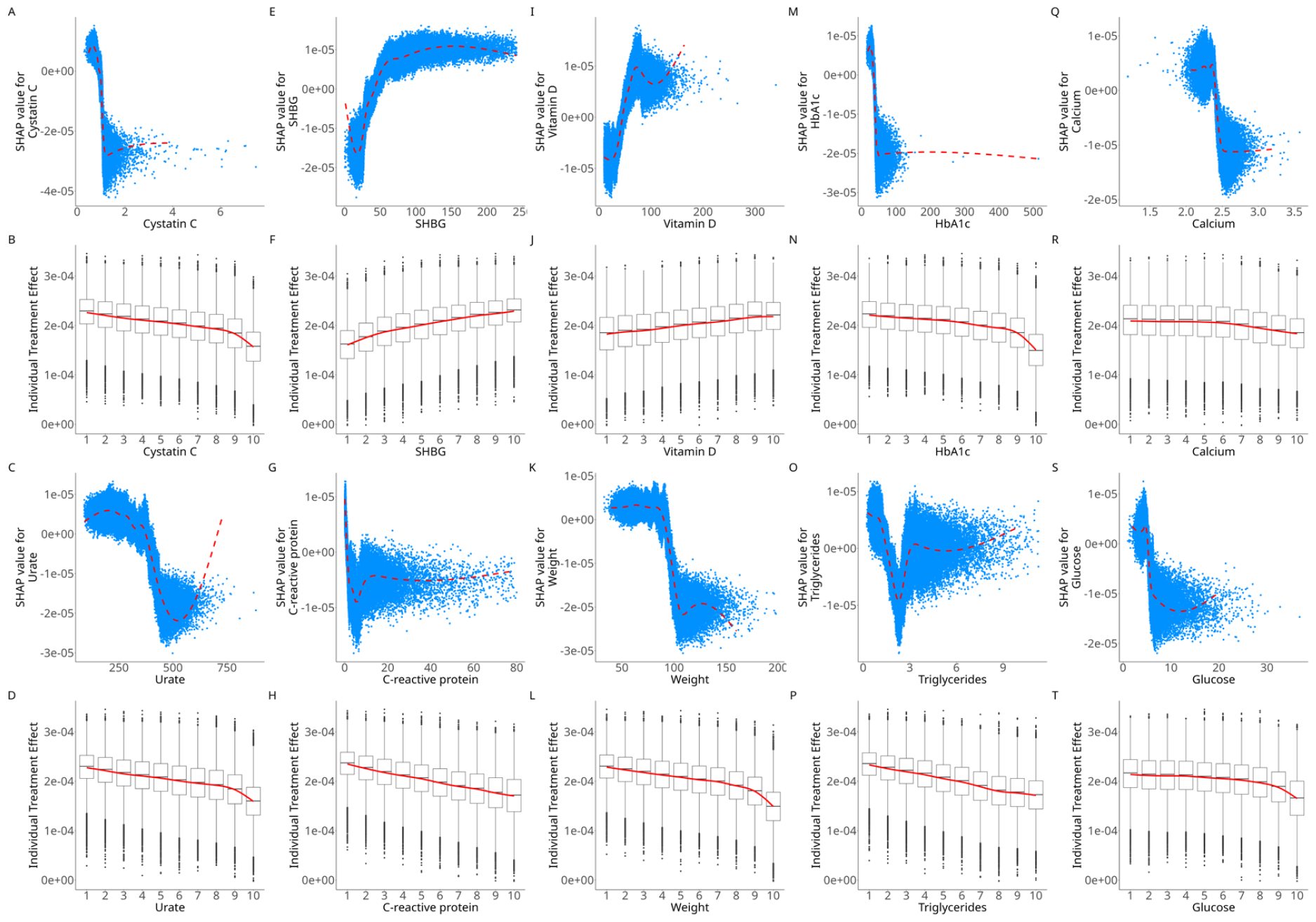
Shapley value plot with Total Cholesterol as risk factor on CAD in continuous treatment setting. A, E, I, M, Q, C, G, K, O, S: Scatterplots of SHAP value (y-axis) versus observed value (x-axis) of top 10 important covariates identified under continuous treatment scenario where Total Cholesterol as risk factor and CAD as the outcome of interest. B, F, J, N, R, D, H, L, P, T: Boxplots of estimated individual treatment effects (y-axis) versus observed value (x-axis) of top 10 important covariates identified under continuous treatment scenario where Total Cholesterol as risk factor and CAD as the outcome of interest.

**Fig 7.**
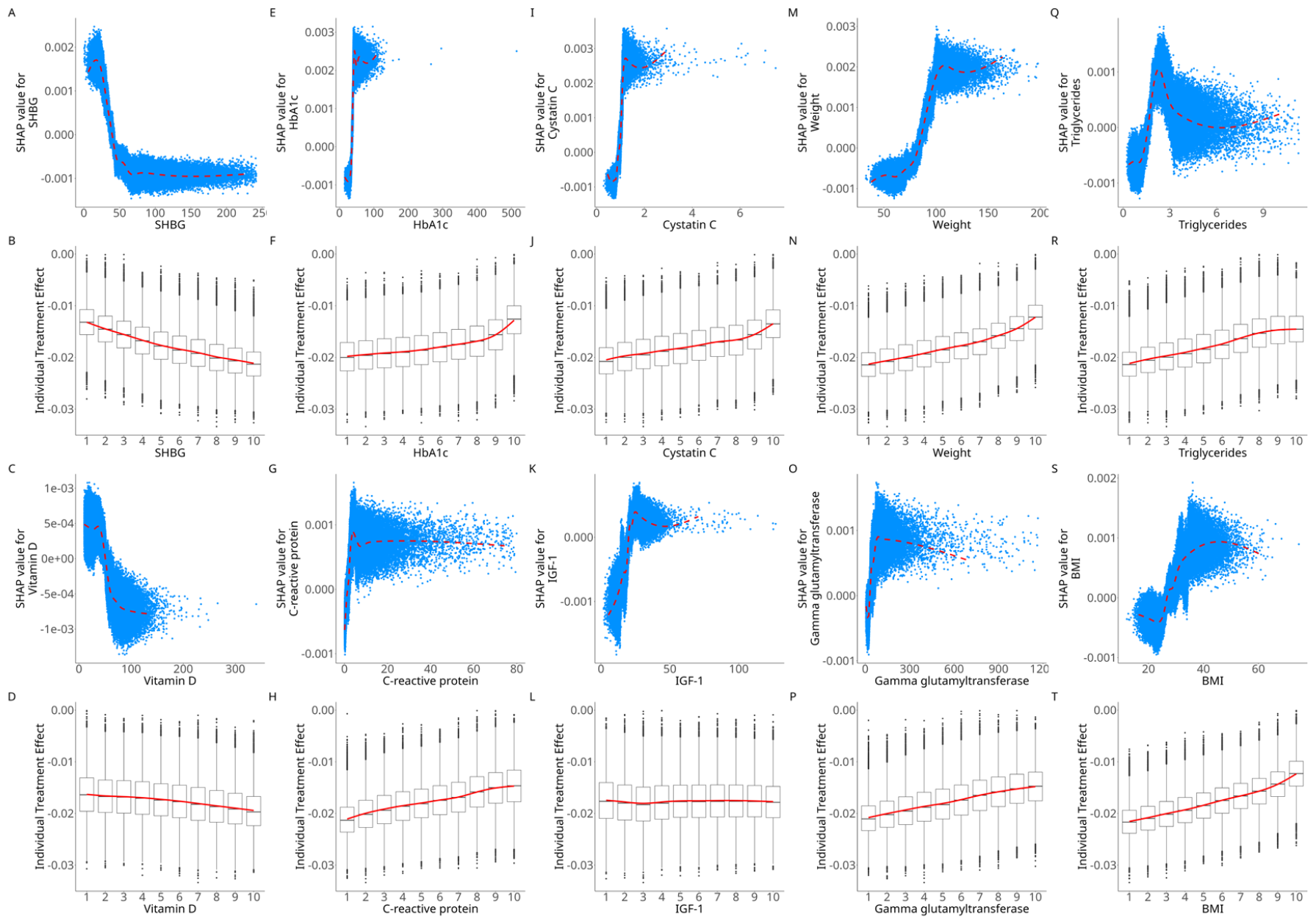
Shapley value plot with Total Cholesterol as risk factor on CAD in binary treatment setting. A, E, I, M, Q, C, G, K, O, S: Scatterplots of SHAP value (y-axis) versus observed value (x-axis) of top 10 important covariates identified under binary treatment scenario where Total Cholesterol as risk factor and CAD as the outcome of interest. B, F, J, N, R, D, H, L, P, T: Boxplots of estimated individual treatment effects (y-axis) versus observed value (x-axis) of top 10 important covariates identified under binary treatment scenario where Total Cholesterol as risk factor and CAD as the outcome of interest.

## 4 Discussion

### 4.1 Overview

In this study, we extend the regular Mendelian randomization approach to infer the *individualized* causal effects of risk factors/treatments in observational studies. Traditional Mendelian randomization primarily focuses on inferring the average causal effect^84^, which may not suffice in the era of precision medicine. Although the estimated average effect is still meaningful in designing policy/treatment for the population, it may obscure individual responses. To address this limitation, we introduce a novel framework that integrates Mendelian Randomization and machine learning methodologies to estimate the individualized treatment effect. Additionally, we present two permutation-based tests to assess the presence of effect heterogeneity within our framework. Through a simulation study, we demonstrate the applicability of our proposed MR framework in practical scenarios likely involving unobserved confounders. Furthermore, we evaluate the performance of our proposed heterogeneity testing approaches through an additional simulation.

As a proof-of-concept example, we applied our framework to study the individualized causal effects of several lipid-related traits on CAD, including LDL-C, HDL-Triglyceride, and Total cholesterol. We found substantial evidence of heterogeneity, particularly for LDL-C and TC’s effect on CAD. Through Shapley value plots, we identified key clinical features that may modify the effects of cholesterol on CAD. This application underscores the utility of our approach and could have significant clinical implications.

This study uncovered important insights that could help optimize the management of dyslipidemia and reduce CAD risk. By characterizing heterogeneous treatment effects, we identified patients who may experience a more pronounced adverse impact of dyslipidemia on CAD risk. These high-risk individuals may be prioritized for intensive lifestyle and pharmacological interventions aimed at lowering lipids.

Targeting treatments to those predicted to derive the greatest risk reduction from lipid control could maximize the efficiency of limited healthcare resources. Moreover, our analysis revealed certain clinical factors associated with varied responses to lipid-modifying therapies. Understanding such sources of treatment heterogeneity can guide clinical decision-making and more personalized prescription. For instance, we have shown that patients who are obese may benefit less from lipid control; weight control in addition to lipid-modifying drugs may lead to more pronounced benefit in terms of CAD prevention.

### 4.2 Strengths and Limitations

Our study possesses several notable strengths. To the best of our knowledge, this is the first study aiming to estimate individualized causal treatment effects leveraging the principles of MR. Although it is possible for researchers to study ITE under an RCT setting, the inherent difficulties and substantial costs associated with designing and implementing an RCT often make such designs impractical. Our approach offers an alternative, enabling the inference of individualized treatment effects using genetic instruments. This method is considerably less susceptible to unknown confounders and reverse causality. This key advantage could further expedite the progress of precision medicine, as interventions on risk factors can be customized for each individual based on the predicted ITE. Furthermore, our ITE estimation strategies are predicated on Machine Learning (ML) methods, which allow flexible modeling of complex relationships. In addition, the DRIV approach also allows virtually any ML model to be used, thereby enhancing the flexibility and applicability of our approach.

Moreover, our simulation results demonstrate that our proposed heterogeneity testing methods exhibited commendable performance in the majority of scenarios. This substantiates that our proposed permutation-based test offers a flexible and robust mechanism for detecting the presence of heterogeneity. Lastly, the integration of SHAP analysis within our framework aids in identifying the primary variables contributing to ITEs. This not only facilitates more comprehensive model explanations but also potentially assists in patient sub-grouping or sub-typing in practical applications.

Our study does present several limitations. For instance, our simulation results show that the permutation variance test may not perform optimally in complex scenarios, such as those involving strong nonlinear treatment effects or interactions between different types of confounders. Additionally, our framework currently only works for a one-sample design since the two stage least square (2SLS) is the fundamental algorithm of our framework in estimating the causal effect between exposure and outcome. Our study primarily focuses on estimating Absolute Risk Reduction (ARR); the study of ITE in terms of Risk Ratio (RR) will be considered as a topic for future studies. Another limitation is the absence of an external dataset for validating our results. It is relatively challenging to find a large-sample, phenotype-rich dataset with genotype information akin to the UK Biobank. Despite these limitations, the estimated Individual Treatment Effects (ITEs) are reasonable, and their range aligns with estimates from previous Randomized Controlled Trials (RCTs) involving lipid-lowering agents. Lastly, our framework primarily considers a linear effect of exposure, even though nonlinear causal effects may be prevalent in practical scenarios ^85^.

### 4.3 Conclusions

In conclusion, we have developed a novel Mendelian Randomization (MR) framework that is capable of estimating individualized causal effects within observational study settings. We have estimated the ITEs of lipid traits on CAD, and have unveiled important clinical features that contribute to effect heterogeneity through Shapley value analyses. It is our hope that our work will pioneer a new research direction for MR studies, providing a novel method for identifying ITEs. Ultimately, we anticipate that these insights will be translated into clinical practice, aiding in the design of more personalized treatment plans for patients.

## Data Availability

The UK Biobank data is available to all registered researchers upon application. All other data produced in the present work are contained in the manuscript.

https://drive.google.com/drive/folders/1LHPVxCOFzzs1jr3wuIanFvpOOtE4c30t?usp=drive_link

## Code Availability

All analysis code can be found in the following GitHub link: https://github.com/yujias424/MR-ITE

## Acknowledgements

This work was supported partially by a National Natural Science Foundation of China grant (NSFC; grant number 81971706), the Lo Kwee Seong Biomedical Research Fund from The Chinese University of Hong Kong and the KIZ-CUHK Joint Laboratory of Bioresources and Molecular Research of Common Diseases, Kunming Institute of Zoology and The Chinese University of Hong Kong, China. An earlier version of this manuscript was posted on ResearchGate in August 2022 (https://www.researchgate.net/publication/362554069_A_framework_for_detecting_causal_effects_of_risk_factors_at_an_individual_level_based_on_principles_of_Mendelian_randomization_Applications_to_cardiovascular_medicine). We would also like to thank Dr Kai Zhao and Ms Alexandria LAU for useful discussions.

## Appendix

Scenario 1:

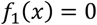

Scenario 2:

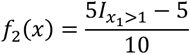

Scenario 3:

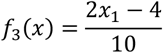

Scenario 4:

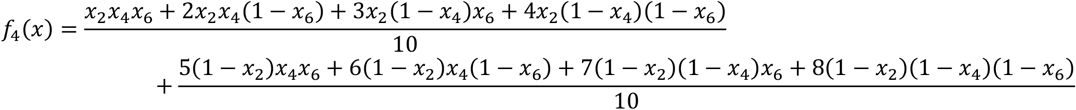

Scenario 5:

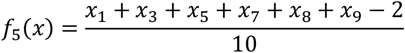

Scenario 6:

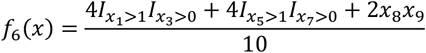

Scenario 7:

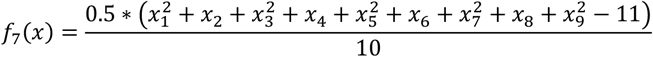

Scenario 8:

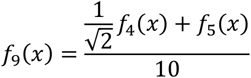

